# Waning of mRNA-1273 vaccine effectiveness against SARS-CoV-2 infection in Qatar

**DOI:** 10.1101/2021.12.16.21267902

**Authors:** Laith J. Abu-Raddad, Hiam Chemaitelly, Houssein H. Ayoub, Hadi M. Yassine, Fatiha M. Benslimane, Hebah A. Al Khatib, Patrick Tang, Mohammad R. Hasan, Peter Coyle, Zaina Al Kanaani, Einas Al Kuwari, Andrew Jeremijenko, Anvar Hassan Kaleeckal, Ali Nizar Latif, Riyazuddin Mohammad Shaik, Hanan F. Abdul Rahim, Gheyath K. Nasrallah, Mohamed Ghaith Al Kuwari, Adeel A. Butt, Hamad Eid Al Romaihi, Mohamed H. Al-Thani, Abdullatif Al Khal, Roberto Bertollini

## Abstract

**BACKGROUND:** In early 2021, Qatar launched a mass immunization campaign with Moderna’s mRNA-1273 COVID-19 vaccine. We assessed persistence of real-world mRNA-1273 effectiveness against SARS-CoV-2 infection and against COVID-19 hospitalization and death.

**METHODS:** Effectiveness was estimated using test-negative, case-control study design, between January 1 and December 5, 2021. Effectiveness was estimated against documented infection (a PCR-positive swab, regardless symptoms), and against any severe (acute-care hospitalization), critical (ICU hospitalization), or fatal COVID-19.

**RESULTS:** By December 5, 2021, 2,962 breakthrough infections had been recorded among those who received two mRNA-1273 doses. Of these infections, 19 progressed to severe COVID-19 and 4 to critical, but none to fatal disease. mRNA-1273 effectiveness against infection was negligible for the first two weeks after the first dose, increased to 65.5% (95% CI: 62.7-68.0%) 14 or more days after the first dose, and reached its peak at about 90% in the first three months after the second dose. Effectiveness declined gradually starting from the fourth month after the second dose and was below 50% by the 7^th^ month after the second dose. Effectiveness against severe, critical, or fatal COVID-19 reached its peak at essentially 100% right after the second dose, and there was no evidence for declining effectiveness over time. Effectiveness against symptomatic versus asymptomatic infection demonstrated the same pattern of waning, but effectiveness against symptomatic infection was consistently higher than that against asymptomatic infection and waned more slowly.

**CONCLUSIONS:** mRNA-1273-induced protection against infection appears to wane month by month after the second dose. Meanwhile, protection against hospitalization and death appears robust with no evidence for waning for several months after the second dose.

## Introduction

In early 2021, Qatar launched a mass immunization campaign with Moderna’s mRNA-1273^1^ Coronavirus Disease 2019 (COVID-19) vaccine.^2^ We assessed persistence of real-world mRNA-1273 effectiveness against severe acute respiratory syndrome coronavirus 2 (SARS-CoV-2) infection and against COVID-19 hospitalization and death.

## Methods

### Study population, data sources, and study design

This study was conducted in the resident population of Qatar, applying the same methodology that was used recently to assess waning of BNT162b2^6^ vaccine effectiveness in the same population.^3^ A detailed description of this methodology can be found in Chemaitelly *et al*.^3^

COVID-19 laboratory testing, vaccination, clinical infection data, and related demographic details were extracted from the national, federated SARS-CoV-2 databases that include all polymerase chain reaction (PCR) testing, COVID-19 vaccinations, and COVID-19 hospitalizations and deaths in Qatar since the start of the pandemic.

Every PCR test conducted in Qatar is classified on the basis of symptoms and the reason for testing (clinical symptoms, contact tracing, surveys or random testing campaigns, individual requests, routine healthcare testing, pre-travel, at port of entry, or other). Qatar has unusually young, diverse demographics, in that only 9% of its residents are ≥50 years of age, and 89% are expatriates from over 150 countries.^7,8^ Nearly all individuals were vaccinated in Qatar, but if vaccinated elsewhere, those vaccinations were still recorded in the health system at the port of entry upon return to Qatar.

Vaccine effectiveness was estimated using the test-negative, case-control study design, a standard design for assessing vaccine effectiveness.^2,9-16^ Cases (PCR-positive persons) and controls (PCR-negative persons) were matched one-to-two by sex, 10-year age group, nationality, reason for SARS-CoV-2 PCR testing, and calendar week of PCR testing to estimate vaccine effectiveness against SARS-CoV-2 infection; and one-to-five to estimate vaccine effectiveness against any severe, critical, or fatal COVID-19 (to improve statistical precision given the relatively small number of severe forms of COVID-19). Matching was performed to control for known differences in the risk of exposure to SARS-CoV-2 infection in Qatar.^8,17-20^

Only the first PCR-positive test during the study was included for each case, and only the first PCR-negative test during the study was included for each control. PCR tests done for pre-travel or at the port of entry were excluded from analysis. All PCR-negative tests for persons included as cases were excluded from analysis. These inclusion and exclusion criteria were implemented to minimize different types of potential bias, as informed by prior analyses.^3^

All persons who received mixed vaccines, or who received a vaccine other than mRNA-1273, or who were tested by PCR after receiving a booster dose were excluded. Every case that met the inclusion criteria and that could be matched to a control was included in the analysis. Both PCR-test outcomes and vaccination status were ascertained at the time of the PCR test.

Effectiveness was estimated against documented infection (defined as a PCR-positive swab, regardless of the reason for PCR testing or the presence of symptoms), as well as against any severe,^21^ critical,^21^ or fatal^22^ COVID-19. Classification of COVID-19 case severity (acute-care hospitalizations),^21^ criticality (ICU hospitalizations),^21^ and fatality^22^ followed World Health Organization (WHO) guidelines, and assessments were made by trained medical personnel using individual chart reviews (Section S1).

Each person who had a positive PCR test result and hospital admission was subject to an infection severity assessment every three days until discharge or death, regardless of the length of the hospital stay or the time between the PCR-positive test and the final disease outcome. Individuals who progressed to severe,^21^ critical,^21^ or fatal^22^ COVID-19 between the PCR-positive test result and the end of the study were classified based on their worst outcome, starting with death, followed by critical disease, and then severe disease.

Details of laboratory methods for real-time reverse-transcription PCR (RT-qPCR) testing are found in Section S2. All PCR testing was conducted at the Hamad Medical Corporation Central Laboratory or at Sidra Medicine Laboratory, following standardized protocols.

The study was approved by the Hamad Medical Corporation and Weill Cornell Medicine-Qatar Institutional Review Boards with a waiver of informed consent. Reporting of the study followed STROBE guidelines (Table S1).

### Statistical analysis

All records of PCR testing in Qatar during the study were included, but only samples of matched cases and controls were included in the analysis. Demographic characteristics of study samples were described using frequency distributions and measures of central tendency.

The odds ratio, comparing odds of vaccination among cases versus controls, and its associated 95% confidence interval (CI) were derived using conditional logistic regression, that is factoring the matching in the study design. This matching and analysis approach aims to minimize potential bias due to variation in epidemic phase,^9,23^ gradual roll-out of vaccination during the study,^9,23^ or other confounders.^24,25^ CIs were not adjusted for multiplicity. Interactions were not investigated. Vaccine effectiveness at different time points and its associated 95% CI were then calculated by applying the following equation.^9,10^

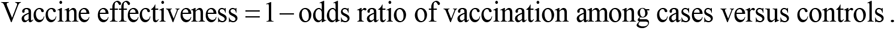

In each analysis for a specific time-since-vaccination stratum, we included only those vaccinated in that specific time-since-vaccination stratum and those unvaccinated (our reference group). Only matched pairs of PCR-positive and PCR-negative persons, in which members of the pair were either unvaccinated or fell within each time-since-vaccination stratum were included in the corresponding vaccine effectiveness estimate. Thus, the number of cases (and controls) varied across time-since-vaccination analyses. Effectiveness after the second dose was estimated month by month, where one month was defined as 30 days.

A sensitivity analysis was conducted by adjusting in the conditional logistic regression for prior infection and healthcare worker status, as healthcare workers were prioritized for vaccination and may have had a different risk of exposure to the infection. The analysis specifically adjusted for being a healthcare worker at Hamad Medical Corporation, the main public healthcare provider in Qatar and the nationally designated provider for all COVID-19 healthcare needs.

Vaccine effectiveness was also estimated against symptomatic infection, defined as a PCR-positive test conducted because of clinical suspicion due to presence of symptoms compatible with a respiratory tract infection, and against asymptomatic infection, defined as a PCR-positive test conducted with no reported presence of symptoms compatible with a respiratory tract infection. In the latter case, PCR testing was done strictly as part of a survey or a random testing campaign. Vaccine effectiveness was further estimated by age group and for severe forms of COVID-19.

## Results

Between January 1 and December 5, 2021, 893,779 individuals received at least one mRNA-1273 dose and 887,726 completed the two-dose regimen. The median date for the first dose was May 27, 2021, and that for the second dose was June 27, 2021. The median time elapsed between the two doses was 28 days (interquartile range, 28-30 days). By December 5, 2021, 2,962 breakthrough infections had been recorded among those who received two doses. Of these infections, 19 progressed to severe COVID-19 and 4 to critical, but none to fatal disease.

Figure 1 depicts the process used to select the study population. Tables 1-3 show the characteristics of samples used in the analysis. Only ∼35% of cases were diagnosed because of symptoms. The remaining cases were diagnosed because of PCR testing for contact tracing, surveys or random testing campaigns, individual requests, and routine healthcare testing.

**Figure 1.**
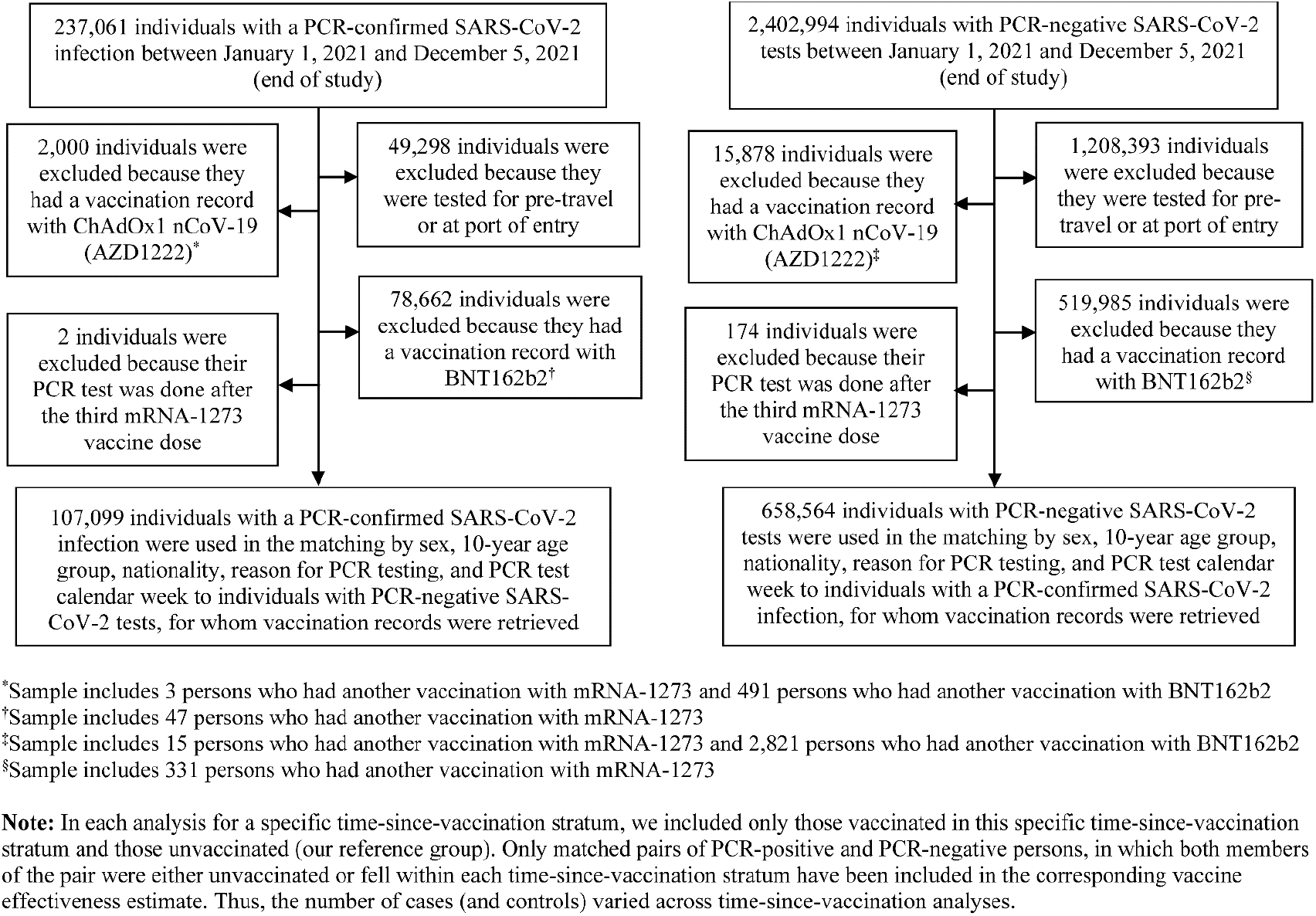
Flowchart describing the population selection process for investigating mRNA-1273 vaccine effectiveness.

**Table 1.** Demographic characteristics of subjects and reasons for PCR testing among samples used to estimate mRNA-1273 vaccine effectiveness. The table includes samples used in the 0-13-days-after-first-dose analysis, ≥14-days-after-first-dose-and-no-second-dose analysis, and 1st-month-after-second-dose analysis.

| Characteristics | 0-13-days-after-first-dose |  |  | ≥14-days-after-first-dose analyses |  |  | 1<sup>st</sup>-month-after-second-dose |  |  |
| --- | --- | --- | --- | --- | --- | --- | --- | --- | --- |
|  | Cases*<br>(PCR-positive) | Controls*<br>(PCR-negative) | SMD<sup>§</sup> | Cases*<br>(PCR-positive) | Controls*<br>(PCR-negative) | SMD<sup>§</sup> | Cases*<br>(PCR-positive) | Controls*<br>(PCR-negative) | SMD<sup>§</sup> |
|  | N=89,767 | N=151,005 |  | N=88,192 | N=149,086 |  | N=87,509 | N=148,349 |  |
| **Median age (IQR) — years** | 30 (16-38) | 30 (12-38) | 0.05<sup>‡</sup> | 30 (15-38) | 30 (12-37) | 0.04<sup>‡</sup> | 30 (14-38) | 30 (11-37) | 0.04<sup>‡</sup> |
| **Age group — no. (%)** |  |  |  |  |  |  |  |  |  |
| <20 years | 23,670 (26.4) | 41,895 (27.7) |  | 23,668 (26.8) | 41,909 (28.1) |  | 23,654 (27.0) | 41,872 (28.2) |  |
| 20-29 years | 18,567 (20.7) | 31,719 (21.0) |  | 18,230 (20.7) | 31,284 (21.0) |  | 18,146 (20.7) | 31,186 (21.0) |  |
| 30-39 years | 28,060 (31.3) | 46,707 (30.9) |  | 27,439 (31.1) | 45,975 (30.8) |  | 27,165 (31.0) | 45,678 (30.8) |  |
| 40-49 years | 14,264 (15.9) | 22,752 (15.1) | 0.04 | 13,771 (15.6) | 22,144 (14.9) | 0.04 | 13,570 (15.5) | 21,936 (14.8) | 0.04 |
| 50-59 years | 4,221 (4.7) | 6,508 (4.3) |  | 4,117 (4.7) | 6,379 (4.3) |  | 4,022 (4.6) | 6,282 (4.2) |  |
| 60-69 years | 769 (0.9) | 1,109 (0.7) |  | 748 (0.9) | 1,083 (0.7) |  | 738 (0.8) | 1,080 (0.7) |  |
| 70+ years | 216 (0.2) | 315 (0.2) |  | 219 (0.3) | 312 (0.2) |  | 214 (0.2) | 315 (0.2) |  |
| **Sex** |  |  |  |  |  |  |  |  |  |
| Male | 61,688 (68.7) | 104,433 (69.2) | 0.01 | 60,377 (68.5) | 102,808 (69.0) | 0.01 | 59,947 (68.5) | 102,396 (69.0) | 0.01 |
| Female | 28,079 (31.3) | 46,572 (30.8) |  | 27,815 (31.5) | 46,278 (31.0) |  | 27,562 (31.5) | 45,953 (31.0) |  |
| **Nationality<sup>†</sup>** |  |  |  |  |  |  |  |  |  |
| Bangladeshi | 6,578 (7.3) | 10,774 (7.1) |  | 6,392 (7.3) | 10,569 (7.1) |  | 6,360 (7.3) | 10,528 (7.1) |  |
| Egyptian | 5,677 (6.3) | 9,271 (6.1) |  | 5,592 (6.3) | 9,197 (6.2) |  | 5,550 (6.3) | 9,162 (6.2) |  |
| Filipino | 8,532 (9.5) | 13,684 (9.1) |  | 8,277 (9.4) | 13,298 (8.9) |  | 8,147 (9.3) | 13,123 (8.9) |  |
| Indian | 24,372 (27.2) | 42,010 (27.8) |  | 23,876 (27.1) | 41,418 (27.8) |  | 23,665 (27.0) | 41,262 (27.8) |  |
| Nepalese | 7,735 (8.6) | 13,248 (8.8) | 0.04 | 7,567 (8.6) | 13,024 (8.7) | 0.04 | 7,531 (8.6) | 12,958 (8.7) | 0.04 |
| Pakistani | 4,960 (5.5) | 8,592 (5.7) |  | 4,908 (5.6) | 8,529 (5.7) |  | 4,881 (5.6) | 8,520 (5.7) |  |
| Qatari | 11,961 (13.3) | 21,413 (14.2) |  | 11,968 (13.6) | 21,437 (14.4) |  | 11,912 (13.6) | 21,364 (14.4) |  |
| Sri Lankan | 3,059 (3.4) | 4,926 (3.3) |  | 2,992 (3.4) | 4,814 (3.2) |  | 2,968 (3.4) | 4,799 (3.2) |  |
| Sudanese | 2,408 (2.7) | 3,875 (2.6) |  | 2,363 (2.7) | 3,823 (2.6) |  | 2,325 (2.7) | 3,777 (2.6) |  |
| Other nationalities<sup>‡</sup> | 14,485 (16.1) | 23,212 (15.4) |  | 14,257 (16.2) | 22,977 (15.4) |  | 14,170 (16.2) | 22,856 (15.4) |  |
| **Reason for PCR testing** |  |  |  |  |  |  |  |  |  |
| Clinical suspicion | 31,238 (34.8) | 43,476 (28.8) |  | 30,736 (34.9) | 43,165 (29.0) |  | 30,381 (34.7) | 42,973 (29.0) |  |
| Contact tracing | 15,077 (16.8) | 26,243 (17.4) |  | 14,834 (16.8) | 26,008 (17.4) |  | 14,663 (16.8) | 25,650 (17.3) |  |
| Healthcare routine testing | 10,999 (12.3) | 19,493 (12.9) | 0.14 | 10,845 (12.3) | 19,329 (13.0) | 0.13 | 10,821 (12.4) | 19,296 (13.0) | 0.13 |
| Survey | 21,676 (24.2) | 41,258 (27.3) |  | 21,195 (24.0) | 40,400 (27.1) |  | 21,026 (24.0) | 40,136 (27.1) |  |
| Individual request | 10,469 (11.7) | 20,115 (13.3) |  | 10,278 (11.7) | 19,774 (13.3) |  | 10,304 (11.8) | 19,874 (13.4) |  |
| Other | 308 (0.3) | 420 (0.3) |  | 304 (0.3) | 410 (0.3) |  | 314 (0.4) | 420 (0.3) |  |

Abbreviations: IQR, interquartile range; PCR, polymerase chain reaction; SMD, standardized mean difference.
<sup>†</sup>Cases and controls were matched one-to-two by sex, 10-year age group, nationality, reason for PCR testing, and calendar week of PCR test.
<sup>‡</sup>Nationalities were chosen to represent the most populous groups in Qatar.
<sup>‡</sup>These comprise 102 other nationalities in Qatar in the 0-13-days-after-first-dose analysis, 102 other nationalities in the ≥14-days-after-first-dose-and-no-second-dose analysis, and 102 other nationalities in the 1st-month-after-second-dose analysis.
<sup>§</sup>SMD is the difference in the mean of a covariate between groups divided by the pooled standard deviation. An SMD<0.1 indicates adequate matching.
<sup>§</sup>SMD is the difference in the mean of a covariate between groups divided by the pooled standard deviation.

**Table 2.** Demographic characteristics of subjects and reasons for PCR testing among samples used to estimate mRNA-1273 vaccine effectiveness. The table includes samples used in the 2^nd^-month-after-second-dose analysis, 3^rd^-month-after-second-dose analysis, and 4^th^-month-after-second-dose analysis.

| Characteristics | 2<sup>nd</sup>-month-after-second-dose |  |  | 3<sup>rd</sup>-month-after-second-dose |  |  | 4<sup>th</sup>-month-after-second-dose |  |  |
| --- | --- | --- | --- | --- | --- | --- | --- | --- | --- |
|  | Cases*<br>(PCR-positive) | Controls*<br>(PCR-negative) | SMD<sup>§</sup> | Cases*<br>(PCR-positive) | Controls*<br>(PCR-negative) | SMD<sup>§</sup> | Cases*<br>(PCR-positive) | Controls*<br>(PCR-negative) | SMD<sup>§</sup> |
|  | N=86,957 | N=147,065 |  | N=86,903 | N=146,880 |  | N=86,976 | N=146,959 |  |
| **Median age (IQR) — years** | 30 (14-38) | 30 (11-37) | 0.04<sup>†</sup> | 30 (11-37) | 30 (14-38) | 0.04<sup>†</sup> | 30 (14-38) | 30 (11-37) | 0.04<sup>†</sup> |
| **Age group — no. (%)** |  |  |  |  |  |  |  |  |  |
| <20 years | 23,653 (27.2) | 41,874 (28.5) |  | 23,639 (27.2) | 41,855 (28.5) |  | 23,642 (27.2) | 41,869 (28.5) |  |
| 20-29 years | 18,051 (20.8) | 30,884 (21.0) |  | 18,023 (20.7) | 30,861 (21.0) |  | 18,032 (20.7) | 30,869 (21.0) |  |
| 30-39 years | 26,976 (31.0) | 45,264 (30.8) |  | 26,979 (31.0) | 45,178 (30.8) |  | 27,013 (31.1) | 45,213 (30.8) |  |
| 40-49 years | 13,408 (15.4) | 21,557 (14.7) | 0.04 | 13,415 (15.4) | 21,543 (14.7) | 0.04 | 13,426 (15.4) | 21,555 (14.7) | 0.04 |
| 50-59 years | 3,935 (4.5) | 6,129 (4.2) |  | 3,917 (4.5) | 6,093 (4.2) |  | 3,931 (4.5) | 6,101 (4.2) |  |
| 60-69 years | 724 (0.8) | 1,050 (0.7) |  | 721 (0.8) | 1,047 (0.7) |  | 724 (0.8) | 1,049 (0.7) |  |
| 70+ years | 210 (0.2) | 307 (0.2) |  | 209 (0.2) | 303 (0.2) |  | 208 (0.2) | 303 (0.2) |  |
| **Sex** |  |  |  |  |  |  |  |  |  |
| Male | 59,526 (68.5) | 101,364 (68.9) | 0.01 | 59,479 (68.4) | 101,225 (68.9) | 0.01 | 59,513 (68.4) | 101,223 (68.9) | 0.01 |
| Female | 27,431 (31.6) | 45,701 (31.1) |  | 27,424 (31.6) | 45,655 (31.1) |  | 27,463 (31.6) | 45,736 (31.1) |  |
| **Nationality<sup>‡</sup>** |  |  |  |  |  |  |  |  |  |
| Bangladeshi | 6,316 (7.3) | 10,408 (7.1) |  | 6,291 (7.2) | 10,357 (7.1) |  | 6,294 (7.2) | 10,358 (7.1) |  |
| Egyptian | 5,507 (6.3) | 9,070 (6.2) |  | 5,505 (6.3) | 9,063 (6.2) |  | 5,512 (6.3) | 9,079 (6.2) |  |
| Filipino | 8,126 (9.3) | 13,065 (8.9) |  | 8,111 (9.3) | 13,022 (8.9) |  | 8,128 (9.4) | 13,080 (8.9) |  |
| Indian | 23,462 (27.0) | 40,739 (27.7) |  | 23,458 (27.0) | 40,674 (27.7) |  | 23,455 (27.0) | 40,645 (27.7) |  |
| Nepalese | 7,507 (8.6) | 12,914 (8.8) | 0.04 | 7,497 (8.6) | 12,882 (8.8) | 0.04 | 7,481 (8.6) | 12,856 (8.8) | 0.04 |
| Pakistani | 4,857 (5.6) | 8,440 (5.7) |  | 4,859 (5.6) | 8,469 (5.8) |  | 4,859 (5.6) | 8,444 (5.8) |  |
| Qatari | 11,839 (13.6) | 21,224 (14.4) |  | 11,823 (13.6) | 21,191 (14.4) |  | 11,861 (13.6) | 21,254 (14.5) |  |
| Sri Lankan | 2,943 (3.4) | 4,753 (3.2) |  | 2,939 (3.4) | 4,741 (3.2) |  | 2,952 (3.4) | 4,751 (3.2) |  |
| Sudanese | 2,313 (2.7) | 3,752 (2.6) |  | 2,317 (2.7) | 3,753 (2.6) |  | 2,318 (2.7) | 3,762 (2.6) |  |
| Other nationalities<sup>‡</sup> | 14,087 (16.2) | 22,700 (15.4) |  | 14,103 (16.2) | 22,728 (15.5) |  | 14,116 (16.2) | 22,730 (15.5) |  |
| **Reason for PCR testing** |  |  |  |  |  |  |  |  |  |
| Clinical suspicion | 30,124 (34.6) | 42,421 (28.9) |  | 30,104 (34.6) | 42,310 (28.8) |  | 30,154 (34.7) | 42,373 (28.8) |  |
| Contact tracing | 14,621 (16.8) | 25,535 (17.4) |  | 14,613 (16.8) | 25,507 (17.4) |  | 14,611 (16.8) | 25,496 (17.4) |  |
| Healthcare routine testing | 10,802 (12.4) | 19,252 (13.1) | 0.13 | 10,798 (12.4) | 19,245 (13.1) | 0.13 | 10,801 (12.4) | 19,248 (13.1) | 0.13 |
| Survey | 20,927 (24.1) | 39,894 (27.1) |  | 20,921 (24.1) | 39,867 (27.1) |  | 20,932 (24.1) | 39,892 (27.1) |  |
| Individual request | 10,176 (11.7) | 19,554 (13.3) |  | 10,164 (11.7) | 19,544 (13.3) |  | 10,175 (11.7) | 19,546 (13.3) |  |
| Other | 307 (0.4) | 409 (0.3) |  | 303 (0.4) | 407 (0.3) |  | 303 (0.4) | 404 (0.3) |  |

Abbreviations: IQR, interquartile range; PCR, polymerase chain reaction.
<sup>†</sup>Cases and controls were matched one-to-two by sex, 10-year age group, nationality, reason for PCR testing, and calendar week of PCR test.
<sup>‡</sup>Nationalities were chosen to represent the most populous groups in Qatar.
<sup>§</sup>These comprise 102 other nationalities in Qatar in the 2<sup>nd</sup>-month-after-second-dose analysis, 102 other nationalities in the 3<sup>rd</sup>-month-after-second-dose analysis, and 102 other nationalities in the 4<sup>th</sup>-month-after-second-dose analysis.
<sup>§</sup>SMD is the difference in the mean of a covariate between groups divided by the pooled standard deviation. An SMD<0.1 indicates adequate matching.
<sup>†</sup>SMD is the difference in the mean of a covariate between groups divided by the pooled standard deviation.

**Table 3.** Demographic characteristics of subjects and reasons for PCR testing among samples used to estimate mRNA-1273 vaccine effectiveness. The table includes samples used in the 5th-month-after-second-dose analysis, 6th-month-after-second-dose analysis, 7th-month-after-second-dose, and 8th-month-after-second-dose analysis.

| Characteristics | 5<sup>th</sup>-month-after-second-dose |  |  | 6<sup>th</sup>-month-after-second-dose |  |  | 7<sup>th</sup>-month-after-second-dose |  |  | 8<sup>th</sup>-month-after-second-dose |  |  |
| --- | --- | --- | --- | --- | --- | --- | --- | --- | --- | --- | --- | --- |
|  | Cases*<br>(PCR-positive) | Controls*<br>(PCR-negative) | SMD<sup>§</sup> | Cases*<br>(PCR-positive) | Controls*<br>(PCR-negative) | SMD<sup>§</sup> | Cases*<br>(PCR-positive) | Controls*<br>(PCR-negative) | SMD<sup>§</sup> | Cases*<br>(PCR-positive) | Controls*<br>(PCR-negative) | SMD<sup>§</sup> |
|  | N=86,962 | N=146,870 |  | N=86,814 | N=146,576 |  | N=86,843 | N=146,628 |  | N=86,754 | N=146,435 |  |
| **Median age (IQR)**<br>— years | 30 (14-38) | 30 (11-37) | 0.04<sup>‡</sup> | 30 (14-38) | 30 (11-37) | 0.05<sup>‡</sup> | 30 (14-38) | 30 (11-37) | 0.04<sup>‡</sup> | 30 (14-38) | 30 (11-37) | 0.04<sup>‡</sup> |
| **Age group — no.**<br>(%) |  |  |  |  |  |  |  |  |  |  |  |  |
| <20 years | 23,647 (27.2) | 41,862 (28.5) |  | 23,638 (27.2) | 41,840 (28.5) |  | 23,645 (27.2) | 41,864 (28.6) |  | 23,644 (27.3) | 41,853 (28.6) |  |
| 20-29 years | 18,027 (20.7) | 30,834 (21.0) |  | 17,990 (20.7) | 30,776 (21.0) |  | 17,989 (20.7) | 30,766 (21.0) |  | 17,963 (20.7) | 30,720 (21.0) |  |
| 30-39 years | 26,990 (31.0) | 45,138 (30.7) |  | 26,924 (31.0) | 45,030 (30.7) |  | 26,965 (31.1) | 45,085 (30.8) |  | 26,917 (31.0) | 44,983 (30.7) |  |
| 40-49 years | 13,434 (15.5) | 21,567 (14.7) | 0.04 | 13,403 (15.4) | 21,487 (14.7) | 0.04 | 13,399 (15.4) | 21,486 (14.7) | 0.04 | 13,381 (15.4) | 21,449 (14.7) | 0.04 |
| 50-59 years | 3,929 (4.5) | 6,111 (4.2) |  | 3,927 (4.5) | 6,092 (4.2) |  | 3,916 (4.5) | 6,079 (4.2) |  | 3,915 (4.5) | 6,076 (4.2) |  |
| 60-69 years | 727 (0.8) | 1,055 (0.7) |  | 724 (0.8) | 1,049 (0.7) |  | 720 (0.8) | 1,044 (0.7) |  | 724 (0.8) | 1,049 (0.7) |  |
| 70+ years | 208 (0.2) | 303 (0.2) |  | 208 (0.2) | 302 (0.2) |  | 209 (0.2) | 304 (0.2) |  | 210 (0.2) | 305 (0.2) |  |
| **Sex** |  |  |  |  |  |  |  |  |  |  |  |  |
| Male | 59,513 (68.4) | 101,201 (68.9) | 0.01 | 59,442 (68.5) | 101,056 (68.9) | 0.01 | 59,425 (68.4) | 101,020 (68.9) | 0.01 | 59,386 (68.5) | 100,939 (68.9) | 0.01 |
| Female | 27,449 (31.6) | 45,669 (31.1) |  | 27,372 (31.5) | 45,520 (31.1) |  | 27,418 (31.6) | 45,608 (31.1) |  | 27,368 (31.6) | 45,496 (31.1) |  |
| **Nationality<sup>†</sup>** |  |  |  |  |  |  |  |  |  |  |  |  |
| Bangladeshi | 6,287 (7.2) | 10,347 (7.1) |  | 6,280 (7.2) | 10333 (7.1) |  | 6,281 (7.2) | 10,335 (7.1) |  | 6,278 (7.2) | 10,328 (7.1) |  |
| Egyptian | 5,526 (6.4) | 9,099 (6.2) |  | 5,516 (6.4) | 9080 (6.2) |  | 5,527 (6.4) | 9,099 (6.2) |  | 5,511 (6.4) | 9,062 (6.2) |  |
| Filipino | 8,098 (9.3) | 12,986 (8.8) |  | 8,082 (9.3) | 12953 (8.8) |  | 8,088 (9.3) | 12,962 (8.8) |  | 8,062 (9.3) | 12,901 (8.8) |  |
| Indian | 23,468 (27.0) | 40,673 (27.7) |  | 23,441 (27.0) | 40608 (27.7) |  | 23,467 (27.0) | 40,657 (27.7) |  | 23,439 (27.0) | 40,602 (27.7) |  |
| Nepalese | 7,487 (8.6) | 12,864 (8.8) |  | 7,478 (8.6) | 12842 (8.8) |  | 7,471 (8.6) | 12,818 (8.7) |  | 7,468 (8.6) | 12,810 (8.8) |  |
| Pakistani | 4,860 (5.6) | 8,436 (5.7) | 0.04 | 4,851 (5.6) | 8422 (5.8) | 0.04 | 4,845 (5.6) | 8,405 (5.7) | 0.04 | 4,845 (5.6) | 8,404 (5.7) | 0.04 |
| Qatari | 11,854 (13.6) | 21,239 (14.5) |  | 11,838 (13.6) | 21182 (14.5) |  | 11,827 (13.6) | 21,187 (14.5) |  | 11,829 (13.6) | 21,191 (14.5) |  |
| Sri Lankan | 2,944 (3.4) | 4,737 (3.2) |  | 2,936 (3.4) | 4728 (3.2) |  | 2,929 (3.4) | 4,717 (3.2) |  | 2,929 (3.4) | 4,717 (3.2) |  |
| Sudanese | 2,318 (2.7) | 3,762 (2.6) |  | 2,316 (2.7) | 3754 (2.6) |  | 2,319 (2.7) | 3,762 (2.6) |  | 2,313 (2.7) | 3,750 (2.6) |  |
| Other nationalities<sup>‡</sup> | 14,120 (16.2) | 22,727 (15.5) |  | 14,076 (16.2) | 22674 (15.5) |  | 14,089 (16.2) | 22,686 (15.5) |  | 14,080 (16.2) | 22,670 (15.5) |  |
| **Reason for PCR testing** |  |  |  |  |  |  |  |  |  |  |  |  |
| Clinical suspicion | 30,153 (34.7) | 42,336 (28.8) |  | 30,084 (34.7) | 42178 (28.78) |  | 30,082 (34.6) | 42,175 (28.8) |  | 30,041 (34.6) | 42,077 (28.7) |  |
| Contact tracing | 14,619 (16.8) | 25,501 (17.4) |  | 14,604 (16.8) | 25485 (17.39) |  | 14,603 (16.8) | 25,481 (17.4) |  | 14,599 (16.8) | 25473 (17.4) |  |
| Healthcare routine testing | 10,801 (12.4) | 19,251 (13.1) | 0.13 | 10,800 (12.4) | 19241 (13.13) | 0.13 | 10,790 (12.4) | 19,224 (13.1) | 0.13 | 10,792 (12.4) | 19226 (13.1) | 0.13 |
| Survey | 20,927 (24.1) | 39,865 (27.1) |  | 20,884 (24.1) | 39790 (27.15) |  | 20,919 (24.1) | 39,853 (27.2) |  | 20,885 (24.1) | 39780 (27.2) |  |
| Individual request | 10,156 (11.7) | 19,509 (13.3) |  | 10,143 (11.7) | 19482 (13.29) |  | 10,149 (11.7) | 19,493 (13.3) |  | 10,143 (11.7) | 19484 (13.3) |  |
| Other | 306 (0.4) | 408 (0.3) |  | 299 (0.3) | 400 (0.27) |  | 300 (0.4) | 402 (0.3) |  | 294 (0.3) | 395 (0.3) |  |

Abbreviations: IQR, interquartile range; PCR, polymerase chain reaction.
<sup>†</sup>Cases and controls were matched one-to-two by sex, 10-year age group, nationality, reason for PCR testing, and calendar week of PCR test.
<sup>‡</sup>Nationalities were chosen to represent the most populous groups in Qatar.
<sup>§</sup>These comprise 102 other nationalities in Qatar in the 5th-month-after-second-dose analysis, 102 other nationalities in the 6th-month-after-second-dose analysis, 102 other nationalities in the 7th-month-after-second-dose analysis, and 102 other nationalities in the 8th-month-after-second-dose analysis.
<sup>§</sup>SMD is the difference in the mean of a covariate between groups divided by the pooled standard deviation. An SMD<0.1 indicates adequate matching.
\*SMD is the difference in the mean of a covariate between groups divided by the pooled standard deviation.

mRNA-1273 effectiveness against infection was negligible for the first two weeks after the first dose, increased to 65.5% (95% CI: 62.7-68.0%) 14 or more days after the first dose, and reached its peak at about 90% in the first three months after the second dose (Figure 2 and Table 4). Effectiveness declined gradually starting from the fourth month after the second dose and was below 50% by the 7^th^ month after the second dose. Effectiveness against severe, critical, or fatal COVID-19 reached its peak at essentially 100% right after the second dose, and there was no evidence for declining effectiveness over time. Sensitivity analysis that adjusted for prior infection and healthcare worker status showed same pattern of waning (Table 5).

**Figure 2.**
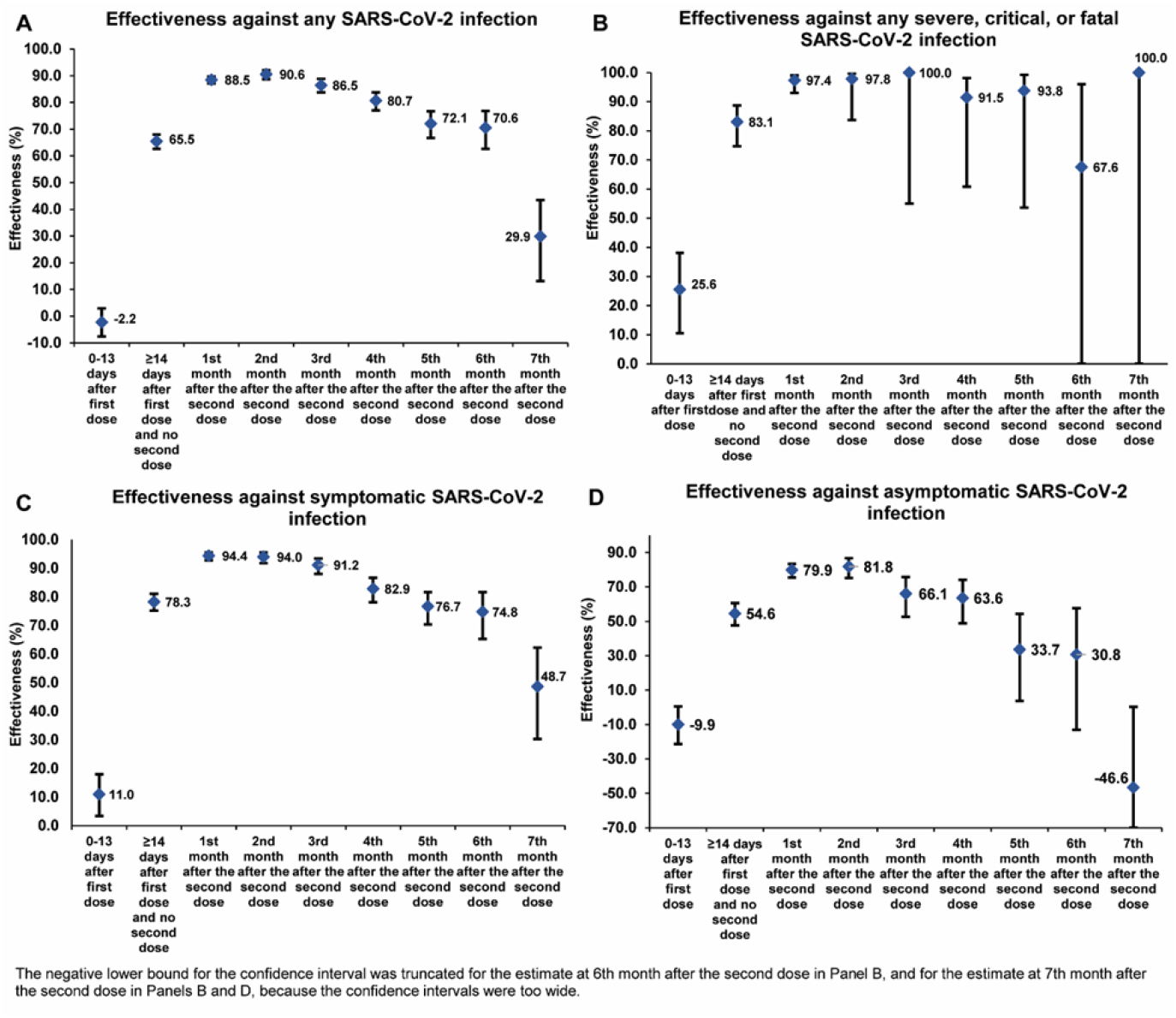
Effectiveness of the mRNA-1273 vaccine against A) any SARS-CoV-2 infection, B) severe, critical, or fatal COVID-19, C) symptomatic SARS-CoV-2 infection, and D) asymptomatic SARS-CoV-2 infection. Data are presented as effectiveness point estimates. Error bars indicate the corresponding 95% confidence intervals.

**Table 4.** Effectiveness of the mRNA-1273 vaccine against any SARS-CoV-2 infection and against any severe, critical, or fatal COVID-19.

| Sub-studies* | Effectiveness against infection |  |  |  |  | Effectiveness against hospitalization and death |  |  |  |  |
| --- | --- | --- | --- | --- | --- | --- | --- | --- | --- | --- |
|  | Cases<sup>†</sup><br>(PCR-positive) |  | Controls<sup>†</sup><br>(PCR-negative) |  | Effectiveness<br>in %<br>(95% CI)<sup>‡</sup> | Cases<sup>†</sup><br>(Severe, critical, or fatal disease)<sup>§</sup> |  | Controls<sup>†</sup><br>(PCR-negative) |  | Effectiveness in<br>%<br>(95% CI)<sup>‡</sup> |
|  | Vaccinated | Unvaccinated | Vaccinated | Unvaccinated |  | Vaccinated | Unvaccinated | Vaccinated | Unvaccinated |  |
| 0-13 days after first dose | 2,931 | 86,836 | 4,360 | 146,645 | -2.2<br>(-7.6 to 2.9) | 172 | 3,073 | 727 | 11,575 | 25.6<br>(10.6 to 38.1) |
| ≥14 days after first dose<br>and no second dose | 962 | 87,230 | 3,891 | 145,195 | 65.5<br>(62.7 to 68.0) | 28 | 3,066 | 464 | 11,379 | 83.1<br>(74.7 to 88.7) |
| 1<sup>st</sup> month after the second<br>dose | 416 | 87,093 | 4,701 | 143,648 | 88.5<br>(87.2 to 89.7) | 4 | 3,068 | 414 | 11,359 | 97.4<br>(93.0 to 99.0) |
| 2<sup>nd</sup> month after the<br>second dose | 148 | 86,809 | 2,136 | 144,929 | 90.6<br>(88.7 to 92.1) | 1 | 3,046 | 132 | 11,492 | 97.8<br>(83.7 to 99.7) |
| 3<sup>rd</sup> month after the second<br>dose | 146 | 86,757 | 1,493 | 145,387 | 86.5<br>(83.8 to 88.8) | 0 | 3,049 | 68 | 11,553 | 100.0<br>(55.0 to 100.0)<sup>¶</sup> |
| 4<sup>th</sup> month after the second<br>dose | 186 | 86,790 | 1,221 | 145,738 | 80.7<br>(77.0 to 83.8) | 2 | 3,043 | 52 | 11,556 | 91.5<br>(60.8 to 98.1) |
| 5<sup>th</sup> month after the second<br>dose | 192 | 86,770 | 902 | 145,968 | 72.1<br>(66.7 to 76.7) | 1 | 3,046 | 44 | 11,566 | 93.8<br>(53.6 to 99.2) |
| 6<sup>th</sup> month after the second<br>dose | 105 | 86,709 | 494 | 146,082 | 70.6<br>(62.7 to 76.8) | 1 | 3,042 | 10 | 11,583 | 67.6<br>(-165.6 to 96.0) |
| 7<sup>th</sup> month after the second<br>dose | 168 | 86,675 | 376 | 146,252 | 29.9<br>(13.1 to 43.5) | 0 | 3,041 | 13 | 11,571 | 100.0<br>(-146.7 to 100.0)<sup>¶</sup> |
| 8<sup>th</sup> month or greater after<br>the second dose | 83 | 86,671 | 137 | 146,298 | 3.5<br>(-33.2 to 30.2) | 0 | 3,043 | 11 | 11,581 | 100.0<br>(-194.4 to 100.0)<sup>¶</sup> |

Abbreviations: CI, confidence interval; PCR, polymerase chain reaction.
<sup>†</sup>In each analysis for a specific time-since-vaccination stratum, we included only those vaccinated in this specific time-since-vaccination stratum and those unvaccinated. Only matched pairs of PCR-positive and PCR-negative persons, in which both members of the pair were either unvaccinated or fell within each time-since-vaccination stratum have been included in the corresponding vaccine effectiveness estimate. Thus, the number of cases (and controls) varied across time-since-vaccination analyses.
<sup>‡</sup>Cases and controls were matched one-to-two by sex, 10-year age group, nationality, reason for PCR testing, and calendar week of PCR test in analysis of effectiveness against infection, and one-to-five in analysis of effectiveness against hospitalization and death.
<sup>§</sup>Vaccine effectiveness was estimated using the test-negative, case-control study design.<sup>9,10</sup>
<sup>¶</sup>Severity,<sup>21</sup> criticality,<sup>21</sup> and fatality<sup>22</sup> were defined as per World Health Organization guidelines.
\*Confidence interval could not be estimated using conditional logistic regression because of zero events among those vaccinated. Alternatively, the confidence interval was estimated using the standard error of the crude odds ratio after adding 0.5 to each of the number of vaccinated cases and number of unvaccinated cases.

**Table 5.** Effectiveness of the mRNA-1273 vaccine against any SARS-CoV-2 infection and against any severe, critical, or fatal COVID-19, after adjusting for prior infection and healthcare worker status.

| Sub-studies* | Effectiveness against infection |  |  |  |  | Effectiveness against hospitalization and death |  |  |  |  |
| --- | --- | --- | --- | --- | --- | --- | --- | --- | --- | --- |
|  | Cases†<br>(PCR-positive) |  | Controls†<br>(PCR-negative) |  | Effectiveness<br>in %<br>(95% CI)‡ | Cases†<br>(Severe, critical, or fatal disease)§ |  | Controls†<br>(PCR-negative) |  | Effectiveness in<br>%<br>(95% CI)‡ |
|  | Vaccinated | Unvaccinated | Vaccinated | Unvaccinated |  | Vaccinated | Unvaccinated | Vaccinated | Unvaccinated |  |
| 0-13 days after first dose | 2,931 | 86,836 | 4,360 | 146,645 | -12.2<br>(-18.3 to -6.4) | 172 | 3,073 | 727 | 11,575 | 22.1<br>(6.1 to 35.3) |
| ≥14 days after first dose<br>and no second dose | 962 | 87,230 | 3,891 | 145,195 | 60.3<br>(57.0 to 63.3) | 28 | 3,066 | 464 | 11,379 | 82.1<br>(73.1 to 88.1) |
| 1<sup>st</sup> month after the second<br>dose | 416 | 87,093 | 4,701 | 143,648 | 85.3<br>(83.5 to 86.9) | 4 | 3,068 | 414 | 11,359 | 97.2<br>(92.4 to 99.0) |
| 2<sup>nd</sup> month after the<br>second dose | 148 | 86,809 | 2,136 | 144,929 | 84.7<br>(81.5 to 87.3) | 1 | 3,046 | 132 | 11,492 | 97.4<br>(81.4 to 99.6) |
| 3<sup>rd</sup> month after the second<br>dose | 146 | 86,757 | 1,493 | 145,387 | 75.7<br>(70.5 to 80.1) | 0 | 3,049 | 68 | 11,553 | 100.0<br>(55.0 to 100.0)¶ |
| 4<sup>th</sup> month after the second<br>dose | 186 | 86,790 | 1,221 | 145,738 | 69.1<br>(62.5 to 74.5) | 2 | 3,043 | 52 | 11,556 | 89.8<br>(51.9 to 97.8) |
| 5<sup>th</sup> month after the second<br>dose | 192 | 86,770 | 902 | 145,968 | 53.6<br>(42.9 to 62.3) | 1 | 3,046 | 44 | 11,566 | 94.2<br>(55.0 to 99.2) |
| 6<sup>th</sup> month after the second<br>dose | 105 | 86,709 | 494 | 146,082 | 50.6<br>(34.5 to 62.8) | 1 | 3,042 | 10 | 11,583 | 61.0<br>(-225.5 to 95.3) |
| 7<sup>th</sup> month after the second<br>dose | 168 | 86,675 | 376 | 146,252 | -3.5<br>(-32.3 to 19.1) | 0 | 3,041 | 13 | 11,571 | 100.0<br>(-146.7 to 100.0)¶ |
| 8<sup>th</sup> month or greater after<br>the second dose | 83 | 86,671 | 137 | 146,298 | -29.5<br>(-84.0 to 8.8) | 0 | 3,043 | 11 | 11,581 | 100.0<br>(-194.4 to 100.0)¶ |

Abbreviations: CI, confidence interval; PCR, polymerase chain reaction.
\*In each analysis for a specific time-since-vaccination stratum, we included only those vaccinated in this specific time-since-vaccination stratum and those unvaccinated. Only matched pairs of PCR-positive and PCR-negative persons, in which both members of the pair were either unvaccinated or fell within each time-since-vaccination stratum have been included in the corresponding vaccine effectiveness estimate. Thus, the number of cases (and controls) varied across time-since-vaccination analyses.
†Cases and controls were matched one-to-two by sex, 10-year age group, nationality, reason for PCR testing, and calendar week of PCR test in analysis of effectiveness against infection, and one-to-five in analysis of effectiveness against hospitalization and death.
‡Vaccine effectiveness was estimated using the test-negative, case-control study design.<sup>9,10</sup>
§Severity,<sup>21</sup> criticality,<sup>21</sup> and fatality<sup>22</sup> were defined as per World Health Organization guidelines.
¶Confidence interval could not be estimated using conditional logistic regression because of zero events among those vaccinated. Alternatively, the confidence interval was estimated using the standard error of the crude odds ratio after adding 0.5 to each of the number of vaccinated cases and number of unvaccinated cases.

Effectiveness for those <50 years of age and those ≥50 were similar in absolute value and showed the same pattern of waning (Table 6). Effectiveness against symptomatic versus asymptomatic infection demonstrated the same pattern of waning, but effectiveness against symptomatic infection was consistently higher than that against asymptomatic infection and waned more slowly (Figure 2 and Table 7). The above measures largely reflected effectiveness against the Beta and Delta variants that dominated incidence during the study.^3-5^

**Table 6.**
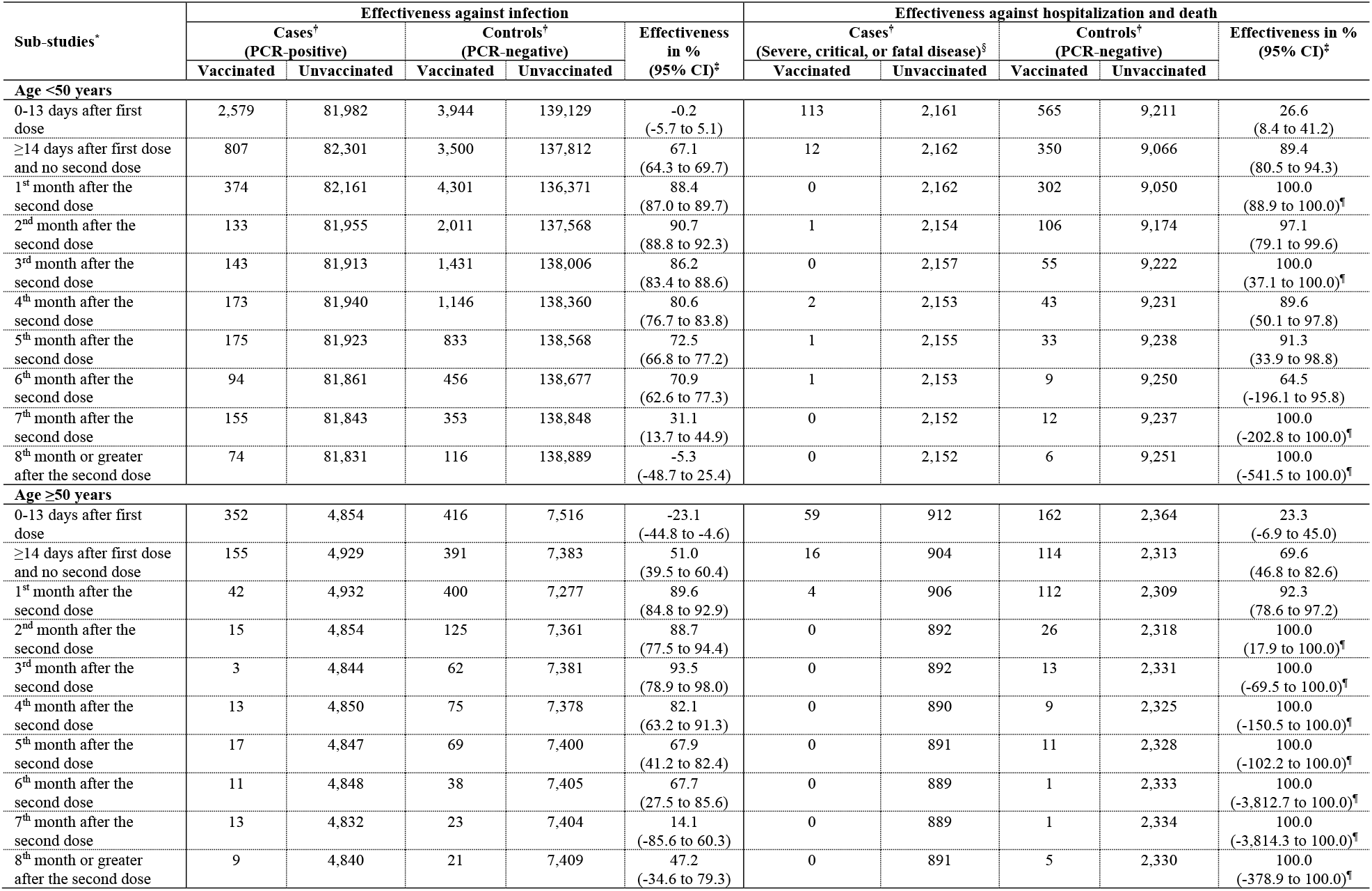
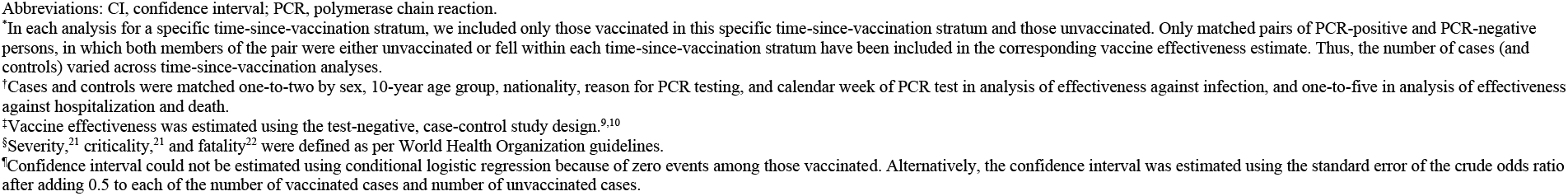
Effectiveness of the mRNA-1273 vaccine against any SARS-CoV-2 infection and against any severe, critical, or fatal COVID-19, stratified by age (<50 years or ≥50 years).

| Sub-studies* | Effectiveness against infection |  |  |  |  | Effectiveness against hospitalization and death |  |  |  |  |
| --- | --- | --- | --- | --- | --- | --- | --- | --- | --- | --- |
|  | Cases<sup>†</sup><br>(PCR-positive) |  | Controls<sup>†</sup><br>(PCR-negative) |  | Effectiveness<br>in %<br>(95% CI)<sup>‡</sup> | Cases<sup>†</sup><br>(Severe, critical, or fatal disease)<sup>§</sup> |  | Controls<sup>†</sup><br>(PCR-negative) |  | Effectiveness in %<br>(95% CI)<sup>‡</sup> |
|  | Vaccinated | Unvaccinated | Vaccinated | Unvaccinated |  | Vaccinated | Unvaccinated | Vaccinated | Unvaccinated |  |
| **Age <50 years** |  |  |  |  |  |  |  |  |  |  |
| 0-13 days after first dose | 2,579 | 81,982 | 3,944 | 139,129 | -0.2<br>(-5.7 to 5.1) | 113 | 2,161 | 565 | 9,211 | 26.6<br>(8.4 to 41.2) |
| ≥14 days after first dose and no second dose | 807 | 82,301 | 3,500 | 137,812 | 67.1<br>(64.3 to 69.7) | 12 | 2,162 | 350 | 9,066 | 89.4<br>(80.5 to 94.3) |
| 1<sup>st</sup> month after the second dose | 374 | 82,161 | 4,301 | 136,371 | 88.4<br>(87.0 to 89.7) | 0 | 2,162 | 302 | 9,050 | 100.0<br>(88.9 to 100.0)<sup>¶</sup> |
| 2<sup>nd</sup> month after the second dose | 133 | 81,955 | 2,011 | 137,568 | 90.7<br>(88.8 to 92.3) | 1 | 2,154 | 106 | 9,174 | 97.1<br>(79.1 to 99.6) |
| 3<sup>rd</sup> month after the second dose | 143 | 81,913 | 1,431 | 138,006 | 86.2<br>(83.4 to 88.6) | 0 | 2,157 | 55 | 9,222 | 100.0<br>(37.1 to 100.0)<sup>¶</sup> |
| 4<sup>th</sup> month after the second dose | 173 | 81,940 | 1,146 | 138,360 | 80.6<br>(76.7 to 83.8) | 2 | 2,153 | 43 | 9,231 | 89.6<br>(50.1 to 97.8) |
| 5<sup>th</sup> month after the second dose | 175 | 81,923 | 833 | 138,568 | 72.5<br>(66.8 to 77.2) | 1 | 2,155 | 33 | 9,238 | 91.3<br>(33.9 to 98.8) |
| 6<sup>th</sup> month after the second dose | 94 | 81,861 | 456 | 138,677 | 70.9<br>(62.6 to 77.3) | 1 | 2,153 | 9 | 9,250 | 64.5<br>(-196.1 to 95.8) |
| 7<sup>th</sup> month after the second dose | 155 | 81,843 | 353 | 138,848 | 31.1<br>(13.7 to 44.9) | 0 | 2,152 | 12 | 9,237 | 100.0<br>(-202.8 to 100.0)<sup>¶</sup> |
| 8<sup>th</sup> month or greater after the second dose | 74 | 81,831 | 116 | 138,889 | -5.3<br>(-48.7 to 25.4) | 0 | 2,152 | 6 | 9,251 | 100.0<br>(-541.5 to 100.0)<sup>¶</sup> |
| **Age ≥50 years** |  |  |  |  |  |  |  |  |  |  |
| 0-13 days after first dose | 352 | 4,854 | 416 | 7,516 | -23.1<br>(-44.8 to -4.6) | 59 | 912 | 162 | 2,364 | 23.3<br>(-6.9 to 45.0) |
| ≥14 days after first dose and no second dose | 155 | 4,929 | 391 | 7,383 | 51.0<br>(39.5 to 60.4) | 16 | 904 | 114 | 2,313 | 69.6<br>(46.8 to 82.6) |
| 1<sup>st</sup> month after the second dose | 42 | 4,932 | 400 | 7,277 | 89.6<br>(84.8 to 92.9) | 4 | 906 | 112 | 2,309 | 92.3<br>(78.6 to 97.2) |
| 2<sup>nd</sup> month after the second dose | 15 | 4,854 | 125 | 7,361 | 88.7<br>(77.5 to 94.4) | 0 | 892 | 26 | 2,318 | 100.0<br>(17.9 to 100.0)<sup>¶</sup> |
| 3<sup>rd</sup> month after the second dose | 3 | 4,844 | 62 | 7,381 | 93.5<br>(78.9 to 98.0) | 0 | 892 | 13 | 2,331 | 100.0<br>(-69.5 to 100.0)<sup>¶</sup> |
| 4<sup>th</sup> month after the second dose | 13 | 4,850 | 75 | 7,378 | 82.1<br>(63.2 to 91.3) | 0 | 890 | 9 | 2,325 | 100.0<br>(-150.5 to 100.0)<sup>¶</sup> |
| 5<sup>th</sup> month after the second dose | 17 | 4,847 | 69 | 7,400 | 67.9<br>(41.2 to 82.4) | 0 | 891 | 11 | 2,328 | 100.0<br>(-102.2 to 100.0)<sup>¶</sup> |
| 6<sup>th</sup> month after the second dose | 11 | 4,848 | 38 | 7,405 | 67.7<br>(27.5 to 85.6) | 0 | 889 | 1 | 2,333 | 100.0<br>(-3,812.7 to 100.0)<sup>¶</sup> |
| 7<sup>th</sup> month after the second dose | 13 | 4,832 | 23 | 7,404 | 14.1<br>(-85.6 to 60.3) | 0 | 889 | 1 | 2,334 | 100.0<br>(-3,814.3 to 100.0)<sup>¶</sup> |
| 8<sup>th</sup> month or greater after the second dose | 9 | 4,840 | 21 | 7,409 | 47.2<br>(-34.6 to 79.3) | 0 | 891 | 5 | 2,330 | 100.0<br>(-378.9 to 100.0)<sup>¶</sup> |

Abbreviations: CI, confidence interval; PCR, polymerase chain reaction.
<sup>a</sup>In each analysis for a specific time-since-vaccination stratum, we included only those vaccinated in this specific time-since-vaccination stratum and those unvaccinated. Only matched pairs of PCR-positive and PCR-negative persons, in which both members of the pair were either unvaccinated or fell within each time-since-vaccination stratum have been included in the corresponding vaccine effectiveness estimate. Thus, the number of cases (and controls) varied across time-since-vaccination analyses.
<sup>c</sup>Cases and controls were matched one-to-two by sex, 10-year age group, nationality, reason for PCR testing, and calendar week of PCR test in analysis of effectiveness against infection, and one-to-five in analysis of effectiveness against hospitalization and death.
<sup>d</sup>Vaccine effectiveness was estimated using the test-negative, case-control study design.<sup>9,10</sup>
<sup>e</sup>Severity,<sup>21</sup> criticality,<sup>21</sup> and fatality<sup>22</sup> were defined as per World Health Organization guidelines.
<sup>f</sup>Confidence interval could not be estimated using conditional logistic regression because of zero events among those vaccinated. Alternatively, the confidence interval was estimated using the standard error of the crude odds ratio after adding 0.5 to each of the number of vaccinated cases and number of unvaccinated cases.

**Table 7.** Effectiveness of the mRNA-1273 vaccine against symptomatic and asymptomatic SARS-CoV-2 infection.

| Sub-studies* | Effectiveness against symptomatic infection† |  |  |  |  | Effectiveness against asymptomatic infection‡ |  |  |  |  |
| --- | --- | --- | --- | --- | --- | --- | --- | --- | --- | --- |
|  | Cases§<br>(PCR-positive) |  | Controls§<br>(PCR-negative) |  | Effectiveness<br>in %<br>(95% CI)¶ | Cases§<br>(PCR-positive) |  | Controls§<br>(PCR-negative) |  | Effectiveness in<br>%<br>(95% CI)¶ |
|  | Vaccinated | Unvaccinated | Vaccinated | Unvaccinated |  | Vaccinated | Unvaccinated | Vaccinated | Vaccinated |  |
| 0-13 days after first dose | 1,170 | 30,068 | 1,562 | 41,914 | 11.0<br>(3.4 to 18.0) | 788 | 20,888 | 1,379 | 39,879 | -9.9<br>(-21.3 to 0.5) |
| ≥14 days after first dose and no<br>second dose | 303 | 30,433 | 1,572 | 41,593 | 78.3<br>(75.2 to 81.1) | 299 | 20,896 | 1,078 | 39,322 | 54.6<br>(47.7 to 60.6) |
| 1<sup>st</sup> month after the second dose | 79 | 30,302 | 1,720 | 41,253 | 94.4<br>(92.8 to 95.6) | 139 | 20,887 | 1,022 | 39,114 | 79.9<br>(75.5 to 83.4) |
| 2<sup>nd</sup> month after the second dose | 44 | 30,080 | 1,033 | 41,388 | 94.0<br>(91.8 to 95.6) | 59 | 20,868 | 432 | 39,462 | 81.8<br>(75.2 to 86.6) |
| 3<sup>rd</sup> month after the second dose | 52 | 30,052 | 814 | 41,496 | 91.2<br>(88.1 to 93.4) | 56 | 20,865 | 241 | 39,626 | 66.1<br>(52.7 to 75.7) |
| 4<sup>th</sup> month after the second dose | 87 | 30,067 | 676 | 41,697 | 82.9<br>(78.1 to 86.7) | 64 | 20,868 | 231 | 39,661 | 63.6<br>(48.8 to 74.2) |
| 5<sup>th</sup> month after the second dose | 94 | 30,059 | 552 | 41,784 | 76.7<br>(70.4 to 81.7) | 63 | 20,864 | 148 | 39,717 | 33.7<br>(3.7 to 54.4) |
| 6<sup>th</sup> month after the second dose | 55 | 30,029 | 301 | 41,877 | 74.8<br>(65.3 to 81.7) | 30 | 20,854 | 73 | 39,717 | 30.8<br>(-13.0 to 57.6) |
| 7<sup>h</sup> month after the second dose | 82 | 30,000 | 225 | 41,950 | 48.7<br>(30.3 to 62.2) | 62 | 20,857 | 86 | 39,767 | -46.6<br>(-115.6 to 0.2) |
| 8<sup>th</sup> month or greater after the<br>second dose | 35 | 30,006 | 64 | 42,013 | 20.0<br>(-29.0 to 50.3) | 29 | 20,856 | 42 | 39,738 | -28.4<br>(-129.3 to 28.1) |

Abbreviations: CI, confidence interval; PCR, polymerase chain reaction.
†In each analysis for a specific time-since-vaccination stratum, we included only those vaccinated in this specific time-since-vaccination stratum and those unvaccinated. Only matched pairs of PCR-positive and PCR-negative persons, in which both members of the pair were either unvaccinated or fell within each time-since-vaccination stratum have been included in the corresponding vaccine effectiveness estimate. Thus, the number of cases (and controls) varied across time-since-vaccination analyses.
‡A symptomatic infection was defined as a PCR-positive test conducted because of clinical suspicion due to presence of symptoms compatible with a respiratory tract infection.
§An asymptomatic infection was defined as a PCR-positive test conducted with no reported presence of symptoms compatible with a respiratory tract infection, that is the PCR testing was done as part of a survey or a random testing campaign.
§Cases and controls were matched one-to-two by sex, 10-year age group, nationality, reason for PCR testing, and calendar week of PCR test in analysis of effectiveness against infection, and one-to-five in analysis of effectiveness against hospitalization and death.
¶Vaccine effectiveness was estimated using the test-negative, case-control study design.<sup>9,10</sup>

## Discussion

mRNA-1273-induced protection against infection appears to wane month by month after the second dose. Meanwhile, protection against hospitalization and death appears robust with no evidence for waning for several months after the second dose.

Since the immunization campaign prioritized vaccination of persons with severe or multiple chronic conditions and by age group, the observed pattern of waning of protection could theoretically be confounded by effects of age and comorbidities. Individual-level data on co-morbid conditions were not available; therefore, they could not be explicitly factored into our analysis. However, only a small proportion of the study population may have had serious co-morbid conditions. Only 9% of the population of Qatar are ≥50 years of age,^7,8^ and 60% are young, expatriate craft and manual workers working in mega-development projects.^19,20,26^ The national list of persons prioritized to receive the vaccine during the first phase of vaccine roll-out included only 19,800 individuals of all age groups with serious co-morbid conditions. Old age may serve as a partial proxy for co-morbid conditions. A similar pattern of waning of protection was observed for younger and older persons (Table 6). Notably, with the small proportion of Qatar’s population being ≥60 years of age,^7,8^ our findings may not be generalizable to other countries in which elderly citizens constitute a larger proportion of the total population.

Infection incidence was dominated sequentially by different variants;^2-5,14,27,28^ thus, it is possible that waning of protection could be confounded by exposure to different variants at different times. However, this seems unlikely, as a similar pattern of waning was observed in our recent study of the BNT162b2 vaccine for the Alpha,^29^ Beta,^29^ and Delta^29^ variants in the same population.^3^

Vaccinated persons presumably have a higher social contact rate than unvaccinated persons, and they may also adhere less strictly to safety measures.^30-32^ This behavior could reduce real-world effectiveness of the vaccine compared to its biological effectiveness, possibly explaining the waning of protection. Public health restrictions have been easing gradually in Qatar, but differently for vaccinated and unvaccinated persons. Many social, work, and travel activities presently require evidence of vaccination (a “health pass”) that is administered through a mandatory mobile app (the Ehteraz app). However, risk compensation is perhaps more likely to affect the overall estimate of effectiveness, rather than the observed waning of protection over time, unless such risk compensation increases with time after the second dose.

PCR testing in Qatar is done on a mass scale, such that about 5% of the population are tested every week.^3^ About 75% of those diagnosed at present are diagnosed not because of symptoms, but because of routine testing.^3^ It is possible that many asymptomatic infections were diagnosed among vaccinated persons that otherwise would have been missed. The higher ascertainment of infection may have reduced the effectiveness estimates. This is supported by the observed lower effectiveness against asymptomatic infection (Table 7).

Effectiveness was assessed using an observational, test-negative, case-control study design,^9,10^ rather than a randomized, clinical trial design, in which cohorts of vaccinated and unvaccinated individuals were followed up. We were unable to use a cohort study design due to depletion of unvaccinated cohorts by the high vaccine coverage. However, the cohort study design applied earlier to the same population of Qatar yielded findings similar to those of the test-negative case-control design,^2,14-16^ supporting the validity of this standard approach in assessing vaccine effectiveness for respiratory tract infections.^2,9-16^ The results of this study are also consistent with our earlier effectiveness estimates immediately after the first and second doses,^2,16^ noting that estimated measures largely reflected effectiveness against the Beta and Delta variants that dominated incidence during that study.^2-5,14,27,28^

To rapidly scale up vaccination, some vaccination campaigns are conducted outside healthcare facilities; thus, records of vaccination are not immediately uploaded into the CERNER system, which tracks all vaccination records at the national level. This administrative time delay can introduce a misclassification bias of those vaccinated versus those unvaccinated. A sensitivity analysis investigating the impact of such potential bias, by assuming a 10% misclassification bias of those vaccinated and unvaccinated in Table 4, found very limited difference in estimated effectiveness. A key strength of the test-negative, case-control study design is that it is less susceptible to this form of bias.^9,10^

Nonetheless, one cannot exclude the possibility that in real-world data, bias could arise in unexpected ways, or from unknown sources, such as subtle differences in test-seeking behavior or changes in the pattern of testing with introduction of other testing modalities, such as rapid antigen testing.

Notwithstanding these limitations, consistent findings were reached, indicating a large effect size for the waning of vaccine protection over time, regardless of the reason for PCR testing, and regardless of the presence or absence of symptoms. Moreover, with the mass scale of PCR testing in Qatar,^3^ the likelihood of bias is perhaps minimized. Extensive sensitivity and additional analyses were conducted to investigate effects of potential bias in our recent study for the BNT162b2 vaccine,^3^ which used the same methodology as the present study. All analyses presented consistent findings of waning vaccine protection.

## Data Availability

The dataset of this study is a property of the Qatar Ministry of Public Health that was provided to the researchers through a restricted-access agreement that prevents sharing the dataset with a third party or publicly. Future access to this dataset can be considered through a direct application for data access to Her Excellency the Minister of Public Health (https://www.moph.gov.qa/english/Pages/default.aspx). Aggregate data are available within the manuscript and its Supplementary information.

## Acknowledgements

We acknowledge the many dedicated individuals at Hamad Medical Corporation, the Ministry of Public Health, the Primary Health Care Corporation, the Qatar Biobank, Sidra Medicine, and Weill Cornell Medicine – Qatar for their diligent efforts and contributions to make this study possible.

The authors are grateful for support from the Biomedical Research Program and the Biostatistics, Epidemiology, and Biomathematics Research Core, both at Weill Cornell Medicine-Qatar, as well as for support provided by the Ministry of Public Health, Hamad Medical Corporation, and Sidra Medicine. The authors are also grateful for the Qatar Genome Programme and Qatar University Biomedical Research Center for institutional support for the reagents needed for the viral genome sequencing. Statements made herein are solely the responsibility of the authors. The funders of the study had no role in study design, data collection, data analysis, data interpretation, or writing of the article.

## Author contributions

LJA conceived and co-designed the study, led the statistical analyses, and co-wrote the first draft of the article. HC co-designed the study, performed the statistical analyses, and co-wrote the first draft of the article. PT and MRH conducted the multiplex, RT-qPCR variant screening and viral genome sequencing. HY, FMB, and HAK conducted viral genome sequencing. All authors contributed to data collection and acquisition, database development, discussion and interpretation of the results, and to the writing of the manuscript. All authors have read and approved the final manuscript.

## Competing interests

Dr. Butt has received institutional grant funding from Gilead Sciences unrelated to the work presented in this paper. Otherwise we declare no competing interests.

## Supplementary Appendix

### Section S1. COVID-19 severity, criticality, and fatality classification

Severe Coronavirus Disease 2019 (COVID-19) disease was defined per the World health Organization (WHO) classification as a severe acute respiratory syndrome coronavirus 2 (SARS-CoV-2) infected person with “oxygen saturation of <90% on room air, and/or respiratory rate of >30 breaths/minute in adults and children >5 years old (or ≥60 breaths/minute in children <2 months old or ≥50 breaths/minute in children 2-11 months old or ≥40 breaths/minute in children 1–5 years old), and/or signs of severe respiratory distress (accessory muscle use and inability to complete full sentences, and, in children, very severe chest wall indrawing, grunting, central cyanosis, or presence of any other general danger signs)”.^1^ Detailed WHO criteria for classifying SARS-CoV-2 infection severity can be found in the WHO technical report.^1^

Critical COVID-19 disease was defined per WHO classification as a SARS-CoV-2 infected person with “acute respiratory distress syndrome, sepsis, septic shock, or other conditions that would normally require the provision of life sustaining therapies such as mechanical ventilation (invasive or non-invasive) or vasopressor therapy”.^1^ Detailed WHO criteria for classifying SARS-CoV-2 infection criticality can be found in the WHO technical report.^1^

COVID-19 death was defined per WHO classification as “a death resulting from a clinically compatible illness, in a probable or confirmed COVID-19 case, unless there is a clear alternative cause of death that cannot be related to COVID-19 disease (e.g. trauma). There should be no period of complete recovery from COVID-19 between illness and death. A death due to COVID-19 may not be attributed to another disease (e.g. cancer) and should be counted independently of preexisting conditions that are suspected of triggering a severe course of COVID-19”. Detailed WHO criteria for classifying COVID-19 death can be found in the WHO technical report.^2^

### Section S2. Laboratory methods

Nasopharyngeal and/or oropharyngeal swabs were collected for PCR testing and placed in Universal Transport Medium (UTM). Aliquots of UTM were: extracted on a QIAsymphony platform (QIAGEN, USA) and tested with real-time reverse-transcription PCR (RT-qPCR) using TaqPath COVID-19 Combo Kits (Thermo Fisher Scientific, USA) on an ABI 7500 FAST (Thermo Fisher, USA); tested directly on the Cepheid GeneXpert system using the Xpert Xpress SARS-CoV-2 (Cepheid, USA); or loaded directly into a Roche cobas 6800 system and assayed with a cobas SARS-CoV-2 Test (Roche, Switzerland). The first assay targets the viral S, N, and ORF1ab gene regions. The second targets the viral N and E-gene regions, and the third targets the ORF1ab and E-gene regions.

All PCR testing was conducted at the Hamad Medical Corporation Central Laboratory or Sidra Medicine Laboratory, following standardized protocols.

**Table S1.** STROBE checklist for case-control studies.

|  | Item No | Recommendation | Main text page |
| --- | --- | --- | --- |
| Title and abstract | 1 | (a) Indicate the study’s design with a commonly used term in the title or the abstract | Abstract |
|  |  | (b) Provide in the abstract an informative and balanced summary of what was done and what was found | Abstract |
| Introduction |  |  |  |
| Background/rationale | 2 | Explain the scientific background and rationale for the investigation being reported | Introduction |
| Objectives | 3 | State specific objectives, including any prespecified hypotheses | Introduction |
| Methods |  |  |  |
| Study design | 4 | Present key elements of study design | Methods (‘Study population, data sources, and study design’) |
| Setting | 5 | Describe the setting, locations, and relevant dates, including periods of recruitment, exposure, follow-up, and data collection | Methods (‘Study population, data sources, and study design’) & Figure 1 |
| Participants | 6 | (a) Give the eligibility criteria, and the sources and methods of case ascertainment and control selection. Give the rationale for the choice of cases and controls | Methods (‘Study population, data sources, and study design’) & Figure 1 |
|  |  | (b) For matched studies, give matching criteria and the number of controls per case |  |
| Variables | 7 | Clearly define all outcomes, exposures, predictors, potential confounders, and effect modifiers. Give diagnostic criteria, if applicable | Methods (‘Study population, data sources, and study design’ & ‘Statistical analysis’), Supp. Sections S1 & S2, & Figure 1 |
| Data sources/measurement | 8 | For each variable of interest, give sources of data and details of methods of assessment (measurement). Describe comparability of assessment methods if there is more than one group | Methods (‘Study population, data sources, and study design’ & ‘Statistical analysis’) & Tables 1-3 |
| Bias | 9 | Describe any efforts to address potential sources of bias | Methods (‘Study population, data sources, and study design’ & ‘Statistical analysis’) |
| Study size | 10 | Explain how the study size was arrived at | Figure 1 |
| Quantitative variables | 11 | Explain how quantitative variables were handled in the analyses. If applicable, describe which groupings were chosen and why | Methods (‘Study population, data sources, and study design’ & ‘Statistical analysis’) & Tables 1-3 |
| Statistical methods | 12 | (a) Describe all statistical methods, including those used to control for confounding | Methods (‘Statistical analysis’) & Tables 1-3 |
|  |  | (b) Describe any methods used to examine subgroups and interactions | Methods (‘Statistical analysis’) |
|  |  | (c) Explain how missing data were addressed | NA, see Methods (‘Study population, data sources, and study design’) |
|  |  | (d) If applicable, explain how matching of cases and controls was addressed | Methods (‘Study population, data sources, and study design’) |
|  |  | (e) Describe any sensitivity analyses | Methods (‘Statistical analysis’) |
| Results |  |  |  |
| Participants | 13 | (a) Report numbers of individuals at each stage of study—eg numbers potentially eligible, examined for eligibility, confirmed eligible, included in the study, completing follow-up, and analysed | Figure 1 |
|  |  | (b) Give reasons for non-participation at each stage |  |
|  |  | (c) Consider use of a flow diagram |  |
| Descriptive data | 14 | (a) Give characteristics of study participants (eg demographic, clinical, social) and information on exposures and potential confounders | Tables 1-3 |
|  |  | (b) Indicate number of participants with missing data for each variable of interest | NA, see Methods (‘Study population, data sources, and study design’) |
| Outcome data | 15 | Report numbers in each exposure category, or summary measures of exposure | Results, Figure 2, & Tables 4-5 |
| Main results | 16 | (a) Give unadjusted estimates and, if applicable, confounder-adjusted estimates and their precision (eg, 95% confidence interval). Make clear which confounders were adjusted for and why they were included | Results, Figure 2, & Tables 4-5 |
|  |  | (b) Report category boundaries when continuous variables were categorized | Tables 1-3 |
|  |  | (c) If relevant, consider translating estimates of relative risk into absolute risk for a meaningful time period | NA |
| Other analyses | 17 | Report other analyses done—eg analyses of subgroups and interactions, and sensitivity analyses | Results & Tables 6-7 |
| **Discussion** |  |  |  |
| Key results | 18 | Summarise key results with reference to study objectives | Results |
| Limitations | 19 | Discuss limitations of the study, taking into account sources of potential bias or imprecision. Discuss both direction and magnitude of any potential bias | Discussion pages 9-10 |
| Interpretation | 20 | Give a cautious overall interpretation of results considering objectives, limitations, multiplicity of analyses, results from similar studies, and other relevant evidence | Discussion page 10 |
| Generalisability | 21 | Discuss the generalisability (external validity) of the study results | Discussion page 10 |
| **Other information** |  |  |  |
| Funding | 22 | Give the source of funding and the role of the funders for the present study and, if applicable, for the original study on which the present article is based | Acknowledgements-page 11 |

Abbreviations: NA, not applicable; p. page; Supp. Supplementary Appendix.

## Notes

### Competing Interest Statement

The authors have declared no competing interest.

### Author Declarations

The study was approved by the Hamad Medical Corporation and Weill Cornell Medicine-Qatar Institutional Review Boards with a waiver of informed consent.

